# Clinical Characteristics of *EYS*-Associated Retinal Dystrophy in 295 Japanese Patients

**DOI:** 10.1101/2024.12.20.24319375

**Authors:** Yoshito Koyanagi, Yusuke Murakami, Taro Kominami, Masatoshi Fukushima, Kensuke Goto, Satoshi Yokota, Kei Mizobuchi, Go Mawatari, Kaoruko Torii, Yuji Inoue, Junya Ota, Daishi Okuda, Kohta Fujiwara, Hanayo Yamaga, Takahiro Hisai, Mikiko Endo, Hanae Iijima, Tomoko Kaida, Kazunori Miyata, Shuji Nakazaki, Takaaki Hayashi, Yasuhiko Hirami, Masato Akiyama, Chikashi Terao, Yukihide Momozawa, Koh-Hei Sonoda, Koji M Nishiguchi, Yasuhiro Ikeda

**Author notes:** Corresponding author: Yasuhiro Ikeda, MD, PhD, Department of Ophthalmology, Faculty of Medicine, University of Miyazaki 5200 Kihara, Kiyotake, Miyazaki, 889-1692, Japan. **Abbreviations:** IRD, Inherited retinal dystrophy; RP, Retinitis Pigmentosa; *EYS*, Eyes shut homolog; BCVA, best-corrected visual acuity; logMAR, logarithm of the minimum angle of resolution; AR, autosomal recessive; WHO, World Health Organization; MAF, minor allele frequency. **Meeting Presentation:** This study was presented in part at the 62nd Annual Meeting of the Japanese Retina and Vitreous Society, the 128th Annual Meeting of the Japanese Ophthalmological Society and the 24th EURETINA Congress. **Statement for the Supplemental Material:** This article contains additional online material. The following should appear online only: Table S3.

## Abstract

**Objective:** To describe the clinical characteristics of Inherited retinal dystrophy (IRD) caused by *EYS* (*EYS*–RD), the most common causative gene of this disease in the Japanese population.

**Design:** A multicenter retrospective study

**Participants:** 295 patients (143 men and 152 women) with *EYS*–RD registered in the Japan Retinitis Pigmentosa Registry Project at eight Japanese facilities.

**Methods:** We evaluated age at the first visit, duration of observation, age of onset, first symptoms, family history, history of consanguineous marriage, disease type, macular complications, history of cataract surgery, logarithm of the minimum angle of resolution best-corrected visual acuity (logMAR BCVA), and its progression. The mean ± standard deviation or the proportion of each parameter was calculated and compared across different variant levels.

**Main Outcome Measures:** Clinical parameters including age of onset, BCVA, and progression of BCVA.

**Results:** The mean age at the first visit was 45.5 ± 14.9 years, and the mean duration of observation was 7.7 ± 6.2 years. The mean age at disease onset was 25.5 ± 14.7 years. The first symptoms of *EYS*–RD included night blindness (78.5%), visual field impairment (9.6%), and loss of visual acuity (8.0%). Family history and consanguineous marriages accounted for 29.7% and 9.3% of the patients, respectively. Rod-cone dystrophy and cone-rod dystrophy accounted for 96.3% and 2.4% of patients, respectively. The mean logMAR BCVA was 0.33 ± 0.56, and the mean progression was 0.03 ± 0.07 per year. In variant-based analyses, three East Asian–specific pathogenic variants (S1653fs, Y2935X, and G843E) caused 69.7% of Japanese *EYS*–RD patients. In cases with homozygous pathogenic variants, the mean age at onset was 17.9, 27.5, and 26.2 years, and the mean progression of logMAR BCVA was 0.05 ± 0.09, 0.04 ± 0.06, and 0.04 ± 0.05 per year for S1653fs (n = 31), Y2935X (n = 13), and G843E (n = 24), respectively.

**Conclusions:** We described the clinical characteristics of Japanese patients with *EYS*–RD. The clinical differences among major East Asian–specific pathogenic variants indicate the utility of genetic testing in personalized medicine for IRD patients tailored to population characteristics.

---

Retinitis Pigmentosa (RP) is the most common form of hereditary retinal degenerative disease, affecting approximately 1 in 3,000–4,000 people, with an estimated 2.5 million people affected worldwide.^1–4^ This genetic disorder involves the slow, progressive degeneration from rod to cone cells, eventually leading to blindness.^1–4^ In Japan, RP is the second leading cause of blindness.^1–3^ RP is the most frequent type of Inherited retinal dystrophy (IRD) and belongs to a group of Mendelian disorders, typically inherited in autosomal dominant, autosomal recessive (AR), or X-linked patterns.^1–4^ Although several gene-specific therapeutic trials have recently been conducted,^4^ there is currently no established treatment for RP.

In ophthalmology, estimating the time until a patient progresses to visual impairment or blindness is a common concern. However, current clinical guidance on this matter remains limited, particularly in relation to IRD. The main reason for this is the genetic diversity of IRD, which has > 300 causative genes (RP: > 90 genes).^5^ Although several studies have identified clinical differences associated with specific causative genes^6, 7^, the genetic background of IRD has been heterogeneous in many clinical studies, including our previous studies.^8–20^ While it is important to understand the natural history of a population with a homogeneous genetic background, the involvement of specific causative genes in clinical characteristics of RP has been limited to evaluation in a small number of cases and this needs to be validated in a large sample size.

Among the causative genes in RP, *Eyes shut homolog* (*EYS*) has been identified in several representative genetic analysis studies^21–24^ and is the most common AR causative gene associated with IRD in the Japanese population.^25–28^ *EYS* is the largest gene related to the eye and was first reported as a gene causing ARRP in 2008.^29^ It spans approximately 2 Mb of chr6q12 and comprises 44 exons encoding a protein of 3146 amino acids that is predominantly expressed in the retina^30^ and contains 27 epidermal growth factor–like domains and five laminin G–like domains that are highly conserved.^29^ Although *EYS* may play an essential role in photoreceptor morphogenesis,^29^ its functional and structural properties remain incompletely characterized. Additionally, its large size has prevented the development of knockout models, resulting in insufficient evaluation of genotype-phenotype correlations. Therefore, studies involving human subjects are crucial for detailed phenotypic analysis of IRD caused by *EYS* (*EYS*–RD).

In this multicenter study, we sought to clarify the clinical characteristics of *EYS*–RD, which is currently the most frequent causative gene in the Japanese population. Using data from the Japan Retinitis Pigmentosa Registry Project at eight Japanese facilities, we included a large number of RP patients with a homogeneous genetic background—295 patients (143 men and 152 women) with *EYS*–RD—with the aim of evaluating the natural history of *EYS*– RD and establishing standard clinical data for Japanese IRD patients.

## Methods

### Ethics Statements

This study was conducted in accordance with the tenets of the 2013 revision of the Declaration of Helsinki and was approved by the local institutional review board. This study was registered with the University Hospital Medical Information Network. Written informed consent was obtained from all patients before sample testing.

### Study Design and Participants

This multicenter retrospective study included 295 patients (143 men and 152 women) with clinical and genetic diagnoses of *EYS*–RD registered at eight Japanese facilities, namely, Jikei University, Teikyo University, Hamamatsu University School of Medicine, Nagoya University, Kobe City Eye Hospital, Kyushu University, Miyata Eye Hospital, and University of Miyazaki.

### Clinical Examination

We retrospectively obtained clinical information from patients’ medical records. The clinical diagnoses of *EYS*–RD were made by trained ophthalmologists at each facility on the basis of the patient’s history of night blindness, visual field constriction, and/or ring scotoma and the results of comprehensive ophthalmological examinations, including slit-lamp biomicroscopy, fundus photography, electroretinography, and optical coherence tomography. Genetic testing was performed for all patients, and *EYS* was confirmed as the causative gene.

The clinical characteristics [age at the first visit, duration of observation, age of onset, first symptoms, family history, history of consanguineous marriage, disease type, macular complications, history of cataract surgery, logarithm of the minimum angle of resolution best-corrected visual acuity (logMAR BCVA) and its progression] of natural disease progression were evaluated. The mean ± standard deviation or proportion of each parameter was calculated. For cases homozygous for one of the three major pathogenic variants [S1653fs and Y2935X^31–33^, and G843E^28, 34^], we evaluated the clinical findings for each variant.

BCVA was measured using the Landolt decimal VA chart at 5 m or with single Landolt test cards. Values were converted to the logarithm of the minimum angle of resolution (logMAR) for statistical evaluation. The logMAR values for counted fingers, hand motion, light perception, and no light perception were defined as 2.3, 2.6, 2.9, and 3.2, respectively.^35^ The World Health Organization (WHO) definitions of low vision (3/10) and blindness (2/40) were used as the threshold values.^36^ Some patient data have been published in a case report or case series.^31, 33, 37^

### Detection of Pathogenic Variants in *EYS*

Peripheral blood samples and salivary specimens were collected from all patients, and genomic DNA was extracted. Genetic variants were assessed using direct Sanger sequencing of *EYS* gene, examined by multiplex polymerase chain reaction–based target sequencing^21^ or whole-exome sequencing with a target analysis of retinal disease–associated genes.^22^ This was then confirmed by direct Sanger sequencing. Variant classification was performed for all detected variants in *EYS* according to the guidelines of the American College of Medical Genetics and Genomics.^23^ Herein, recently reported novel variants were also determined to be pathogenic variants causing *EYS*–RD. The included variants were annotated using Annovar software (version 3.4)^38^ and SnpEff (version 4.3)^39^. Minor allele frequency (MAF) was obtained from gnomAD v4^40^, 14KJPN^41^, and the Korean Genome Project^42^.

### Statistical Analysis

Comparisons of clinical data between the two groups were performed using Student’s two-tailed t-test. Multigroup comparisons were performed using one-way analysis of variance with Bonferroni’s correction. The association between genetic status and the rate of vision loss was evaluated using age- and sex-adjusted linear regression analyses. The Kruskal– Wallis test was used to analyze between-group differences in the logarithm of the number of viral DNA copies and the ratio of viral reads to other reads. We then estimated age of low vision (logMAR BCVA = 0.523) and blindness (logMAR BCVA = 1.3) using a linear mixed-effects model for repeated measurement data.^20, 43–45^ Only cases with two points data of logMAR BCVA without IOL were included. The model included age and group as fixed effects to estimate the overall effects of aging and group differences on visual acuity, with random intercepts and slopes for each individual to account for differences in baseline visual acuity and the rate of change over time. The unstructured covariance matrix was employed to allow distinct correlations between the repeated measurements within individuals. The model’s parameter estimates were used to estimate the age at which visual acuity would decline to logMAR BCVA = 0.523 and logMAR BCVA = 1.3. The two-sided *P* < 0.05 was considered statistically significant. All statistical analyses were performed using R software (version 3.4.4).

## Results

### Demographic Characteristics

Demographic characteristics of the 295 patients with *EYS*–RD are shown in **Table 1**. The mean age at the first visit was 45.5 ± 14.9 years, mean duration of observation was 7.7 ± 6.2 years, and mean age at disease onset was 25.5 ± 14.7 years. The first symptoms of *EYS*– RD included night blindness (78.5%), visual field impairment (9.6%), and loss of visual acuity (8.0%). Family history and consanguineous marriages accounted for 29.7% and 9.3% of the patients, respectively. Rod-cone dystrophy and cone-rod dystrophy accounted for 96.3% and 2.4% of patients, respectively. In the right eye, 11.5%, 4.1%, and 0.3% of the patients had a history of epiretinal membrane, macular edema, and rhegmatogenous retinal detachment, respectively.

**Table 1.** Demographic characteristics of 295 patients with *EYS*-associated retinal dystrophy.

|  | **All (N = 295)** |
| --- | --- |
| **Mean age ± SD (years) at first visit** | 45.5 ± 14.9 |
| **Gender: women, N (%)** |  |
| Men / Women | 143 (48%) / 152 (52%) |
| **Mean follow-up period (range) (years)** | 7.7 ± 6.2 (0 - 27.4) |
| **Mean age ± SD (years) at onset** | 25.5 ± 14.7 |
| **Symptom at onset** |  |
| Night blindness | 197 (78.5%) |
| Visual field defects | 24 (9.6%) |
| Visual acuity decrease | 20 (8.0%) |
| Others | 10 (4.0%) |
| **Family History** | 87 (29.7%) |
| **Consanguinity of parents** | 27 (9.3%) |
| **Disease type, N (%)** |  |
| Rod-cone dystrophy | 283 (96.3%) |
| Cone-rod dystrophy | 7 (2.4%) |
| Others | 4 (1.4%) |
| **Ocular characteristics** |  |
| **Lens status: pseudophakia, N (%)** | 100 (33.9%) |
| **Previous vitrectomy, N (%)** | 4 (1.4%) |
| **Macula complication** |  |
| Eepiretinal membrane | 34 (11.5%) |
| Macular edema | 12 (4.1%) |
| Rhegmatogenous retinal detachment | 1 (0.3%) |
| **Visual Acuity** |  |
| Mean logMAR BCVA ± SD at first visit | 0.33 ± 0.56 |
| Mean logMAR BCVA decrease ± SD / yr | 0.03 ± 0.07 |

Discrepancies in the total arise due to the presence of missing values within the dataset.

### Pathogenic Variants Causing *EYS*–RD in Japan

A schematic representation of the *EYS* protein domains and the distribution of pathogenic variants is shown in **Figure 1A**. Classifications of the detected pathogenic variant types in *EYS*–RD are shown in **Figure 1B**. In total, 56.5% were missense, 21.3% were frameshifts, 16.4% were nonsense, 3.28% were large structural variants, 1.64% were in-frame deletions, and 0.82% were splice-site variants.

**Figure 1.**
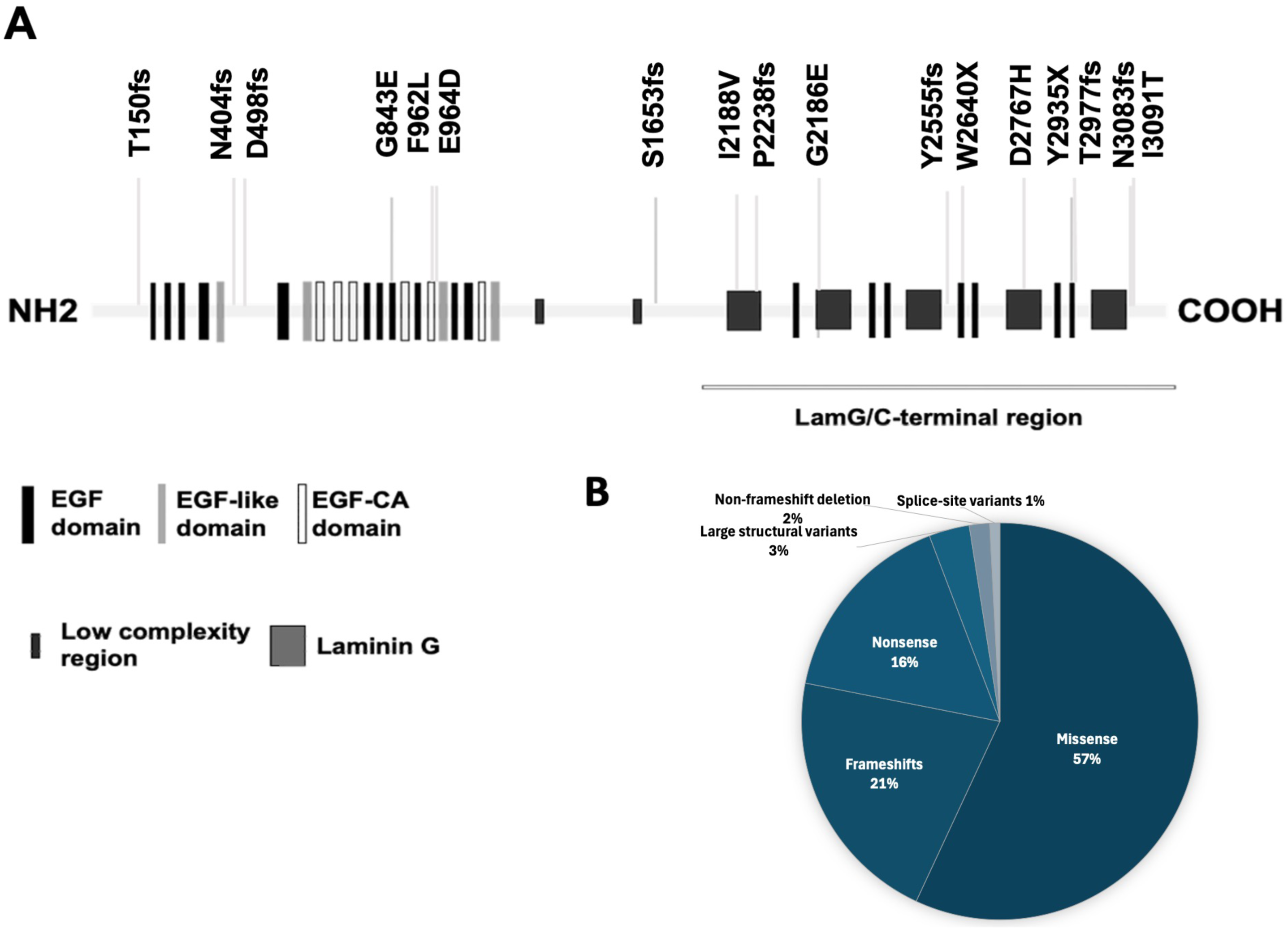
Characteristics of the pathogenic variants causing EYS-associated retinal dystrophy in Japanese patients. (A) Schematic representation of the *EYS* protein domains and the distribution of pathogenic variants; (B) Classifications of the detected pathogenic variant types in EYS-RD.

### Human Lifespan and logMAR BCVA at the First and Last Visit for *EYS*–RD

In 64.4% of patients without cataract surgery, the mean logMAR BCVA at first visit was 0.33 ± 0.56, and the mean progression was 0.03 ± 0.07 per year (**Figure 2**).

**Figure 2.**
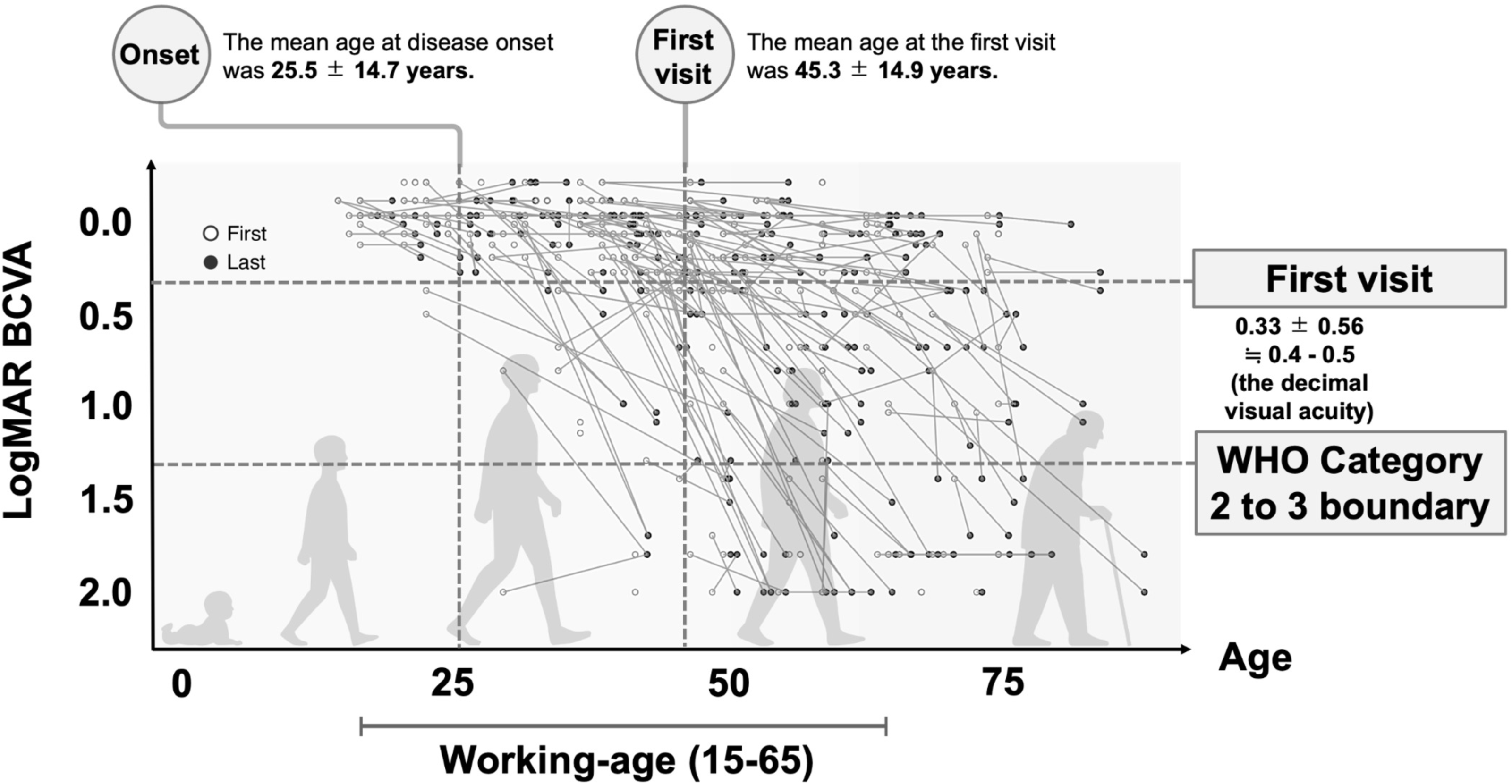
Human lifespan and the scatterplot with paired points of the logMAR visual acuity at the first and last visit in patients with *EYS*-associated retinal dystrophy

Approximately 50% of the patients had low vision (3/10), and 20% were blind (2/40). However, 25% of patients aged ≥65 years maintained a visual acuity of 0.3 or better (decimal BCVA [WHO category 0]). The estimated ages of low vision and blindness (WHO definitions) (95% confidence intervals [CIs]) in *EYS*–RD were 53.9 [47.9, 59.9] and 74.1 [60.4, 87.7] years, respectively.

### Multivariate Analysis of Factors Affecting BCVA

Multivariate analysis of the factors affecting BCVA at the first visit showed that age at the first visit, age at onset, and history of cataract surgery significantly affected visual acuity (**Table 2**). The multivariate evaluation of the factors affecting changes in corrected visual acuity per year showed that two macular complications, namely, macular pseudohole and macular hole, significantly affected the progression of visual acuity loss (**Table S3**).

**Table 2.** Multivariate analysis of factors affecting best corrected visual acuity at first visit.

|  | **Estimate** | **Std. Error** | **P value** |
| --- | --- | --- | --- |
| **Age at first visit (years)** | 0.016 | 0.0052 | 0.0026* |
| **Age at onset (years)** | -0.009 | 0.004 | 0.017* |
| **Gender** | -0.106 | 0.111 | 0.342 |
| **Lens status: pseudophakia** | -1.243 | 0.299 | 0.0002* |
| **Macula complication** |  |  |  |
| Eepiretinal membrane | -0.039 | 0.267 | 0.884 |
| Macular edema | -0.080 | 0.355 | 0.640 |
| Vitreomacular traction syndrome | -0.152 | 0.448 | 0.735 |
| Macular pseudohole | 0.288 | 0.500 | 0.568 |
| Macular hole | -0.427 | 0.461 | 0.360 |
| **Family History** | 0.052 | 0.116 | 0.654 |
| **Consanguinity of parents** | 0.006 | 0.167 | 0.974 |

### Variant-Based Analyses

The frequency of pathogenic AR variant combinations is shown in **Figure 3A**. Approximately seventy percent of these *EYS*–RD patients harbored the three major pathogenic variants (S1653fs, Y2935X, and G843E), and the most frequent combination was S1653fs and G843E, followed by homozygous S1653fs, homozygous G843E, and homozygous Y2935X. As shown in **Figure 3B**, these three major variants were specific to East Asian populations.

**Figure 3.**
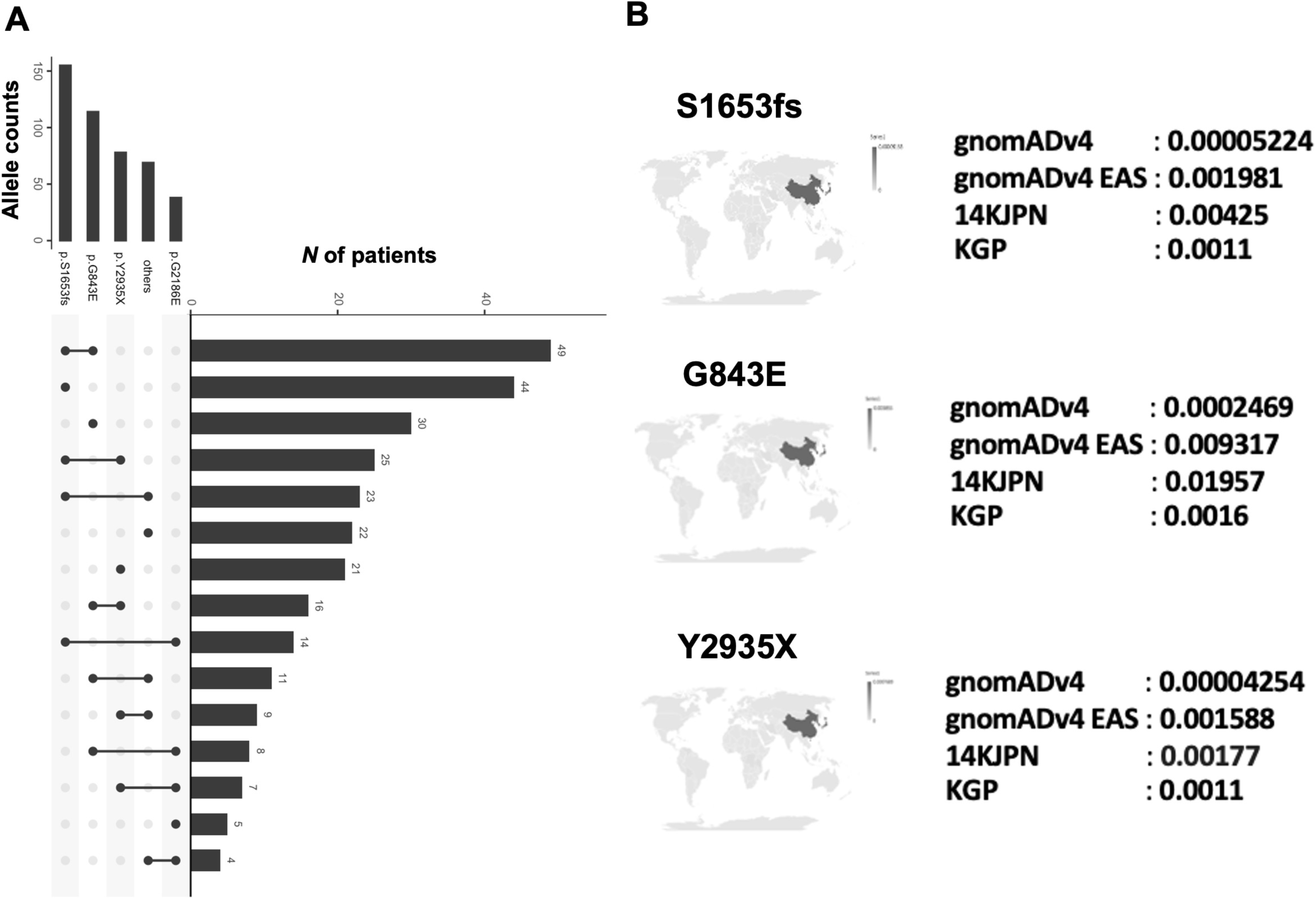
The frequent combinations of the two AR pathogenic variants (A) and world distribution of the top 3 frequent pathogenic variants **(B).**

Comparisons of the clinical characteristics of the homozygotes of the three major pathogenic variants (S1653fs, Y2935X, and G843E) are shown in **Table 4**. The onset of *EYS*–RD caused by homozygous S1653fs occurred earlier than that caused by the other two variants (G843E vs. S1653fs: *P* = 0.016; Y2935X vs. S1653fs: *P* = 0.015) (**Figure 4**). The mean progressions of visual acuity were 0.05 ± 0.09, 0.04 ± 0.05, and 0.04 ± 0.06 per year in S1653fs, G843E, and Y2935X, respectively. Analysis by the linear mixed-effects model revealed that compared to the homozygotes S1653fs, G843E had a non-significant difference in the effect on visual acuity (logMAR BCVA) with a coefficient of -0.074 (95% CI: -0.332, 0.184) (*P* = 0.57). Homozygotes Y2935X, however, showed a statistically significant difference, with a coefficient of -0.284 (95% CI: -0.524, -0.045) (*P* = 0.020). The estimated ages of blindness (95% CIs) in homozygotes of S1653fs, G843E, and Y2935X were 72.0 (58.7, 85.3), 73.9 (59.4, 88.5), 79.5 (63.1, 96.0) years old, respectively, though there was no statistical difference among groups (*P* > 0.05) (**Table 4**).

**Figure 4.**
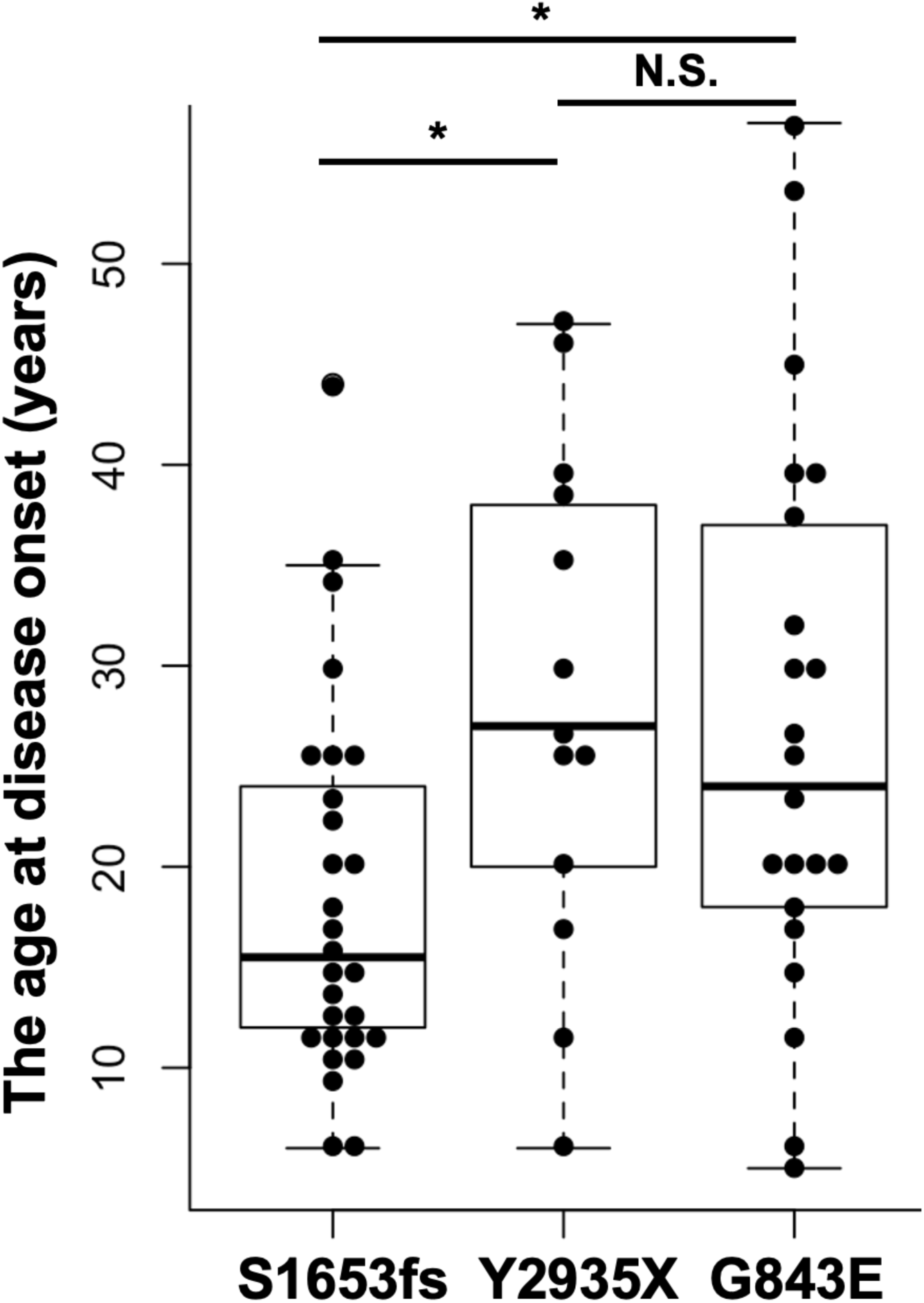
Comparison of the age at disease onset among the homozygotes of the three major pathogenic variants (S1653fs, Y2935X, and G843E). Mean age at disease onset was compared using the Wilcoxon rank-sum test with Bonferroni correction (α= 0.05/3). The single asterisk indicates statistically significant. **N.S.** indicates not significant.

**Table 4.** Comparison of the characteristics of *EYS*-RD among the three major homozygous pathogenic variants.

| **Variant** | **Types** | **N of cases** | **Mean age at disease onset** | **Mean age at the first visit** | **Mean logMAR BCVA at first visit OD** | **Mean progression/yr** | **The estimated age at the decimal BCVA of 0.05 (95% CIs)** |
| --- | --- | --- | --- | --- | --- | --- | --- |
| S1653fs | Frameshift insertion | 31 | 17.9 | 41.7 | 0.34 | 0.05 ± 0.09 | 72.0 (58.7, 85.3) |
| G843E | Missense | 24 | 26.2 | 50.3 | 0.37 | 0.04 ± 0.05 | 73.9 (59.4, 88.5) |
| Y2935X | Stop gain | 13 | 27.5 | 43.2 | 0.01 | 0.04 ± 0.06 | 79.5 (63.1, 96.0) |

## Discussion

In this multicenter retrospective study, we described the clinical characteristics of IRD caused by *EYS* (*EYS*–RD), which is currently the most common causative gene in the Japanese population. Data from 295 patients with *EYS*–RD showed that the mean progression of visual acuity (logMAR BCVA)/year was 0.03 (i.e., difference of a decrease from 1.0 to 0.9 [decimal visual acuity]), indicating that *EYS*–RD may present a mild phenotype of cone dysfunction. We further identified three major genetic variants (S1653fs, Y2935X, and G843E) harboring approximately seventy percent of Japanese *EYS*–RD, all of which were specific to the East Asian population. Comparisons of the clinical characteristics of the homozygotes of these three variants demonstrated that S1653fs may be relatively severe in clinical characteristics of *EYS*–RD.

The strength of this study is that it is the largest *EYS*-RD clinical study based on an estimated 15 million general population. Importantly, most patients (96%) with *EYS*–RD showed rod–cone dystrophy, confirming the fact that *EYS* is the causative gene of IRD. Previous reports in a European (Portuguese) cohort (*N* = 58) of patients with *EYS*–RD showed a similar age of onset and disease type rates.^46^ In addition, the mean ΔlogMAR BCVA of 0.03 per year was comparable to the 1.45–1.51 letters per year (0.028) in the European *EYS*–RD patients.^46^ Given that typical RP shows a progression of 2.3 letters (0.045) per year,^47^ *EYS*–RD may present a mild phenotype of cone dysfunction. Further, the decline in visual acuity generally begins after the age of 40s in this study, and this decline may occur earlier than in the natural history of other IRDs (ex. Bietti’s crystalline dystrophy^48^ or RP caused by S-antigen visual arrestin variants^49^). The period before the decline in visual acuity begins in *EYS*-RD may represent an optimal window for gene therapy and neuroprotective treatments.

While the clinical characteristics of *EYS*-RD are similar to those reported in the previous study in the European patients,^46^ it is noteworthy that the genetic background differs among populations. Specifically, although *EYS*-RD adheres to an autosomal recessive inheritance pattern, this study revealed a low incidence of family history and consanguinity. Instead, we identified the involvement of high-frequency variants specific to East Asia. This finding suggests that the three major pathogenic variants prevalent in the Japanese population may contribute to the sporadic occurrence of *EYS*-RD in a setting of random mating, thereby defining the characteristics of Japanese *EYS*-RD.

Variations in the age of onset and disease progression among three major pathogenic variants may suggest the utility of genetic testing in genetic counseling for IRD patients, including life planning. In particular, the S1653fs-RD patients exhibited an earlier onset compared to patients with the other two high-frequency pathogenic variants (Y2935X and G843E). Moreover, the effect of S1653fs on visual acuity decline was significantly stronger than that of Y2935X. This frameshift insertion variant, located in the middle of the protein structure, was suggested to have a relatively high and severe impact on the phenotype of *EYS*-RD.

This study had several limitations. First, there were difficulties in collecting further clinical data due to interrupted outpatient visits and follow-ups, or failure to collect information despite contact. Specifically, the number of patients aged > 70 years was limited and therefore the progress of this age group could not be adequately assessed. Second, although we performed the largest *EYS*-RD study to date using data from the Japan Retinitis Pigmentosa Registry Project, prospective studies or research conducted in other institutions or East Asian populations (e.g., in China and Korea) are necessary to validate our findings. Third, as for the visual acuity assessment, it was based on evaluations at two time points, the first and last visits, with the analysis assuming linearity in changes for all ages. Fourth, we evaluated the change in BCVA, which means we evaluated cone dysfunction. The analysis of rod function, including visual field loss, is warranted. Furthermore, cases involving structural variants, such as copy number variations (CNVs), large deletions, and Alu insertions, were not well represented in our study. In Japan, the evaluation of structural variants in genetic screening has not been sufficiently conducted. Future studies will need to address the clinical phenotypes of Japanese *EYS*-RD caused by these variants^50–56^.

In conclusion, we described the clinical characteristics of Japanese patients with *EYS*– RD caused by pathogenic variants mainly specific to East Asia. The clinical differences among major East Asian–specific pathogenic variants of *EYS*–RD indicate the utility of genetic testing in personalized medicine for IRD patients tailored to population characteristics.

## Supporting information

coi_disclosure

## Data Availability

All data produced in the present study are available upon reasonable request to the authors.

## Acknowledgments

We acknowledge the staff of the Laboratory for Genotyping Development in RIKEN and the RIKEN-IMS Genome Platform.

