## Supplementary material for "Clinical Characteristics of *EYS*-Associated Retinal Dystrophy in 295 Japanese Patients": Table S3

**Table S3. Factors Affecting Change in best corrected visual acuit per year in multivariate analysis**

| | Estimate | Std. Error | P value |
|---|---|---|---|
| **Age at first visit (years)** | -0.0005 | 0.0011 | 0.637 |
| **Age at onset (years)** | 0.001 | 0.0007 | 0.819 |
| **Gender** | 0.022 | 0.021 | 0.298 |
| **LogMAR BCVA at first visit** | 0.019 | 0.029 | 0.498 |
| **Lens status: pseudophakia** | 0.064 | 0.066 | 0.336 |
| **Macula complication** | | | |
| Eepiretinal membrane | 0.054 | 0.049 | 0.275 |
| Macular edema | 0.234 | 0.088 | 0.155 |
| Vitreomacular traction syndrome | 0.159 | 0.0806 | 0.054 |
| Macular pseudohole | 0.244 | 0.0865 | 0. 0073* |
| Macular hole | 0.178 | 0.083 | 0. 037* |
| **Family History** | -0.007 | 0.022 | 0.748 |
| **Consanguinity of parents** | -0.036 | 0.031 | 0.243 |
